# Adequacy of essential opioid analgesic consumption for anesthesia across 137 countries and territories from 2017 to 2021

**DOI:** 10.1101/2023.05.30.23290676

**Authors:** Shirish Rao, Bhavya Ratan Maroo, Poorvikha Gowda, Siddhesh Zadey

**Affiliations:** Association for Socially Applicable Research (ASAR), Pune, Maharashtra, India; Seth G.S. Medical College and K.E.M. Hospital, Mumbai, Maharashtra, India; Maulana Azad Medical College, Delhi, India; St John’s Medical College, Bangalore, India; Department of Epidemiology, Mailman School of Public Health, Columbia University, NYC NY USA; GEMINI Research Center, Duke University School of Medicine, Durham NC USA; Dr. D. Y. Patil Dental College and Hospital, Dr. D. Y. Patil Vidyapeeth, Pune, Maharashtra, India

**Author notes:** **Correspondence:** Siddhesh Zadey, BSMS MSc-GH; ASAR Office Address: D2 Sai Heritage, New DP Road, Aundh, Pune, Maharashtra, India 411007. **Author Contributions: *Shirish Rao:*** Conceptualization, Methodology, Formal Analysis, Data Curation, Writing - Original Draft, Review & Editing. ***Bhavya Ratan Maroo:*** Methodology, Data Curation, Writing - Review & Editing Poorvika Gowda: Writing - Original Draft, Review & Editing. ***Siddhesh Zadey:*** Conceptualization, Methodology, Formal Analysis, Visualizations, Writing - Review & Editing, Supervision, Project Administration. **Funding:** No funding was received to conduct this study. **Ethics approval and consent to participate:** Not applicable. **Consent for publication:** Not applicable. **Availability of data and materials:** All data used and produced in the manuscript can be accessed from Harvard Dataverse (https://doi.org/10.7910/DVN/F6RFMM).

**Keywords:** Anesthesia, perioperative care, global surgery, opioids, pain management, low- and middle-income countries

## Abstract

**Introduction:** The Adequacy of Opioid Consumption (AOC) Index uses the human development index (HDI) to benchmark pain management. This does not account for health system factors such as the anesthesia workforce and can misrepresent high consumption as better. We improved the AOC index by adjusting it for physician anesthesia provider (PAP) density to provide a better indicator for pain management and perioperative care.

**Methods:** Country-level mean opioid consumption in milligrams per capita (mg/capita) for 2017-2021 (five-year arithmetic mean) was obtained from the International Narcotics Control Board Annual Report 2022. For parsimonious analysis, we included 11 opioids and analogs present in the WHO Model List of Essential Medicines 2023. Projections for PAP density per 100,000 people for 2019 were based on the World Federation of Society of Anaesthesiologists Survey (2015-16) and physician density estimates (2015-2019) derived from the Institute for Health Metrics and Evaluation. A generalized linear regression model for 137 countries was run with mean essential opioid analgesic consumption as the dependent variable and PAP density and HDI (2019) were independent variables to get PAP-adjusted consumption values. The arithmetic mean of PAP-adjusted consumption values of the top 20 countries was used as the adequacy threshold. PAP-adjusted AOC index was calculated as the ratio of the country’s adjusted essential opioid analgesic consumption to the threshold multiplied by 100.

**Results:** PAP-adjusted AOC index values ranged from 129.14 for Switzerland to 0.23 for Mali. Merely 7.3% of countries had a high AOC. About 5.97 billion people are estimated to be living in regions of low to extremely low PAP-adjusted AOC which are mainly situated in low and middle-income countries in the global south.

**Conclusion:** In this comprehensive up-to-date global analysis, we find that most low- and lower-middle-income countries lack access to essential opioid analgesics. These point to the need for investing in the anesthesia workforce and ensuring access to opioid analgesics in tandem.

Research in context

Evidence before this study
Before undertaking the analysis, a rapid review of the literature was conducted to assess existing evidence on global opioid analgesic consumption and adequacy. Relevant publications included comparing opioid consumption for varying numbers of countries using data from the International Narcotics Control Board (INCB), IQVIA MIDAS database, the Organisation for Economic Cooperation and Development, and the European Monitoring Centre for Drugs and Drug Addiction. Three studies quantified the adequacy of opioid consumption between 2006-2015 using ACM and AOC indices.

The added value of this study
Our study provides the latest available data for essential opioid analgesic consumption (190 countries) and adequacy (137 countries) between 2017-2021. We only include opioids and their analogs enlisted in the WHO list of Essential Drugs and use a modified AOC index which is adjusted for Physician Anesthesia Provider (PAP) density. This makes our index specific for anesthesia and perioperative pain management. Adjusting for PAP density normalizes the distribution of AOC by bringing higher and lower extremes closer to the mean adjusted consumption and thus accounts for opioid overconsumption values in developed countries.

Implications of all the available evidence
PAP-adjusted AOC can be used as an indicator to assess regional disparities in access to anesthesia and pain management. AOC has already been used as one of the indicators to measure access to anesthesia and pain management in the South Asian Region. Its national and sub-national mapping can help identify unmet needs and aid in drafting tailored healthcare policies and plans. Further research should be focused on adjusting the AOC based on other health system indicators like surgical rate and burden of disease, and also explore more global indicators for anesthesia care which would aid in tracking its process in global healthcare development.

## Introduction

Access to perioperative care is critical for equitable surgical, obstetric, trauma, and anesthesia (SOTA) care globally.^1^ Prescription opioid analgesics are among the effective pharmacological interventions that are crucial for pain alleviation during and after surgeries.^2^ Inadequate pain control can lead to multiple negative repercussions including delayed post-op recovery, increased risk of infection, and prolonged hospital stays.^3^ Access to opioid analgesics is essential to attenuate the risks associated with pain. Previous studies have assessed the access to opioids in terms of mean annual consumption using data from multiple sources including the International Narcotics Control Bureau (INCB) and IQVIA MIDAS, for data till 2019.^4–7^ However, the mean annual consumption does not directly notify adequacy, as it doesn’t account for health system-based needs. The adequacy of essential opioid analgesic consumption can fill this gap as it can be an indicator proxying safe anesthesia delivery in the broader global SOTA care discourse.

In 2006, Seya et al. developed the Adequacy of Consumption Measure (ACM), a morbidity-corrected measure indicating adequacy based on a country’s opioid consumption, morbidity, and Human Development Index (HDI).^8^ An ACM of 1 or higher was considered adequate.^8^ Duthey and Scholten later updated the ACM by expressing it as a percentage where 100% considered higher adequacy.^4^ In 2019, the measure was further modified and named the Adequacy of Opioid Consumption (AOC) index by Scholten et al.^9^ This study provided global adequacy values until 2015. The AOC index used only the country’s opioid consumption level and HDI to set normative adequacy levels. Morbidity corrections for cancer, HIV, and injuries were excluded from the AOC Index due to problems concerning the unreliability and limited availability of the required data. However, this index discounts health system factors which are essential to quantify the need-based adequacy of opioid analgesics. Incorporating health system factors like Physician Anesthesia Density (PAP) can help adjust the AOC Index by regulating the consumption of prescription opioids with respect to workforce density.

In this analysis, using the latest global opioid consumption dataset for the year 2017-2021 we formulated an updated AOC Index adjusted for PAP density to provide a better indicator for pain management and perioperative care.

## Methods

We conducted a cross-sectional retrospective analysis. Data for opioid consumption for 2017-2021 was obtained from the International Narcotics Control Board (INCB) Annual Report 2022.^10^ From this dataset, only the opioids included in the WHO Model List of Essential Medicines 2023 and their analogs were included in the analysis.^11^ These opioid analgesics were Codeine, Dihydro-codeine, Ethyl-morphine, Hydrocodone, Morphine, Oxycodone, Methadone, Fentanyl, Alfentanil, Remifentanil, and Sufentanil. Data of 197 countries and territories, for each year was obtained in kilograms (kg) and was converted into milligrams per capita using UN Populations Projections of those particular years.^12^ We then calculated the 5-year arithmetic mean of essential opioid analgesic consumption (mg/capita). Countries that had data for at least one year were included in the analysis (190 countries), however, seven countries had no data for any of the 5 years, so they were excluded from the analysis. The HDI data of included countries for the year 2019 (the central year for 2017-2021), was obtained from the UN Human Development Report 2019.^13^

The World Federation of Societies of Anaesthesiologists (WFSA) Global Anesthesia Workforce Survey provided PAP density (per 10,000 people) data for 153 countries and territories for the year 2015-16.^14^ To make all the variables uniform for the year 2019, PAP density projections for 2019 were calculated. For this, we used physician density estimates along with their 95% uncertainty values, for 2015-2019 for 204 countries which were derived from the Institute for Health Metrics and Evaluation (IHME). Using these two data sets, we calculated the ratio of PAP to total physician density (per 100,000 population) for 2016 **(Equation 1)**.^15^

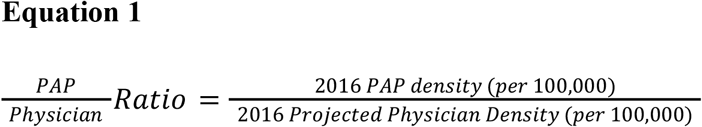

We assumed a uniform scale-up such that the PAP/Physician ratio would remain constant over the years and calculated the 2019 PAP density (per 100,000) by multiplying the PAP/Physician ratio (2016) with 2019 physician density (per 100,000) projections **(Equation 2)**. On similar lines, 95% uncertainty values for 2019 PAP density (per 100,000) were calculated by multiplying the PAP/Physician ratio with 2019 physician density (per 100,000) projections 95% uncertainty values. These projections were validated using recently published data for Anesthetist densities for 2019 for 19 countries by Bouchard et al.^16^

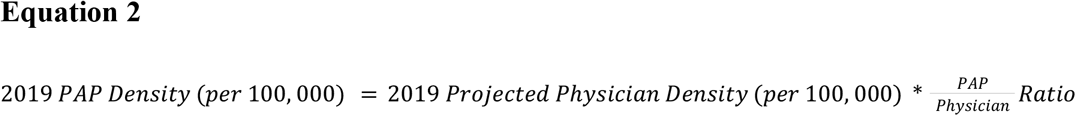

After accounting for country-wise missing data for all three variables i.e. mean essential opioid analgesic consumption (mg/capita) HDI 2019 and 2019 PAP density (per 100,000), complete data for all three variables was available for 137 countries and territories. The population of these 137 countries and territories is representative of 95.3% of the global population as of 2019.

To calculate the PAP-adjusted AOC index, we used a generalized linear regression model. Since the individual distribution of both mean essential opioid analgesic consumption (mg/capita) and PAP density 2019 (per 100,000) were found to be highly skewed towards zero (using Shapiro-Wilk’s test), their natural log was calculated to normalize their distribution. In the regression analysis of log-transformed variables, mean essential opioid analgesic consumption was taken as the dependent variable whereas PAP density 2019 and HDI 2019 were taken as independent variables. Within the regression model, the marginal effect of HDI on opioid analgesic consumption was analyzed. This was used to understand the PAP-adjusted effect of HDI on opioid analgesic consumption. In the regression equation, regression coefficients of HDI and PAP density were substituted, maintaining a constant mean PAP density value **(Equation 3)**. This equation was used to predict the PAP-adjusted essential opioid consumption values. Once the predicted values for PAP-adjusted opioid analgesic consumption were calculated, all the values were back-transformed from Log. Further, these PAP-adjusted opioid analgesic consumption values were used to calculate the PAP-adjusted AOC threshold. This was done by calculating the arithmetic mean of PAP-adjusted opioid analgesic consumption values of the top twenty countries **(Equation 4)**. It is important to note that we have taken the values of the top twenty adjusted consumption countries and not the top twenty HDI countries as HDI has already been accounted for in the regression model. Country-wise PAP-adjusted AOC index was calculated as the country’s PAP-adjusted opioid analgesic consumption divided by the threshold and the fraction multiplied by a hundred **(Equation 5)**.

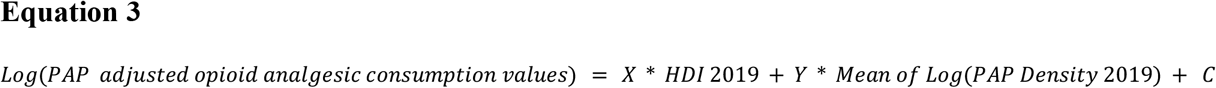

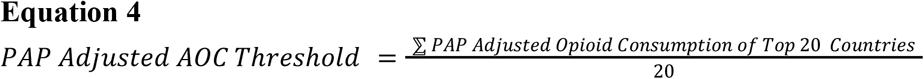

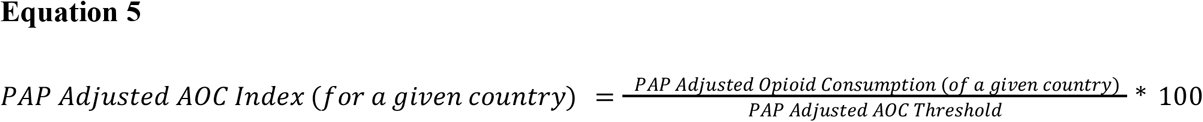

The original AOC threshold was calculated by taking the arithmetic mean of the mean essential opioid analgesic consumption (non-adjusted; as per INCB) of the top twenty HDI countries (Equation 6). The original AOC index was calculated as the ratio of the country’s mean essential opioid analgesic consumption to the threshold multiplied by 100 (Equation 7).

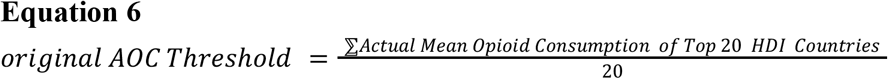

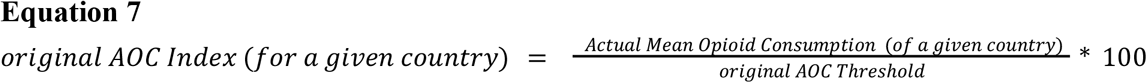

Originally, the AOC index is classified as adequate (AOC >100), moderate (< 100 and >30), low (< 30 and >10), very low (< 10 and >3), and extremely low (< 3). These levels were created as equidistant points on the Logarithmic scale. We kept the cutoffs the same but changed “Adequate” to “High”, as the absence of an upper limit may lead to misinterpretation of the consumption levels which are much higher than 100 as adequate. The number of people living in regions of each PAP-adjusted AOC category was calculated by pooling the population of countries falling under each adequacy category. PAP-adjusted AOC index was also calculated for the World Bank Income Groups (WBIGs) and WHO regions. Its values across WBIGs and WHO regions were compared using Welch’s analysis of variance tests. Spearman rank correlation was used to check for similarities and differences in the distribution of original and PAP-adjusted AOC index values. Further, adequacy categories (based on cut-offs) were compared for original and adjusted methods to check for the number of countries, WBIGs, and WHO regions whose categories changed. A significance level of 5% was used for all tests. Analysis was conducted in Google Sheets, JASP (0.17.2.1), and RStudio. All data created and used in this study as well as the analysis models are available in Harvard Dataverse and **Supplements 1** respectively.^17^

## Results

From 2017-2021, the mean essential opioid analgesic consumption globally (190 countries) was 227.59 mg/capita. The mean annual consumption changed from 249.49 in 2017, 246.24 in 2018, 214.18 in 2019, 218.09 in 2020 and 209.92 in 2021. Among countries and territories for which data from all 5 years was available, Gibraltar (2232.05 mg/capita) (a British overseas territory) had the highest mean consumption whereas mean consumption was as low as 0.17mg/capita in Burkina Faso **(Figure 1a)**. Among countries, Iceland (1645.93 mg/capita) had the highest mean consumption.

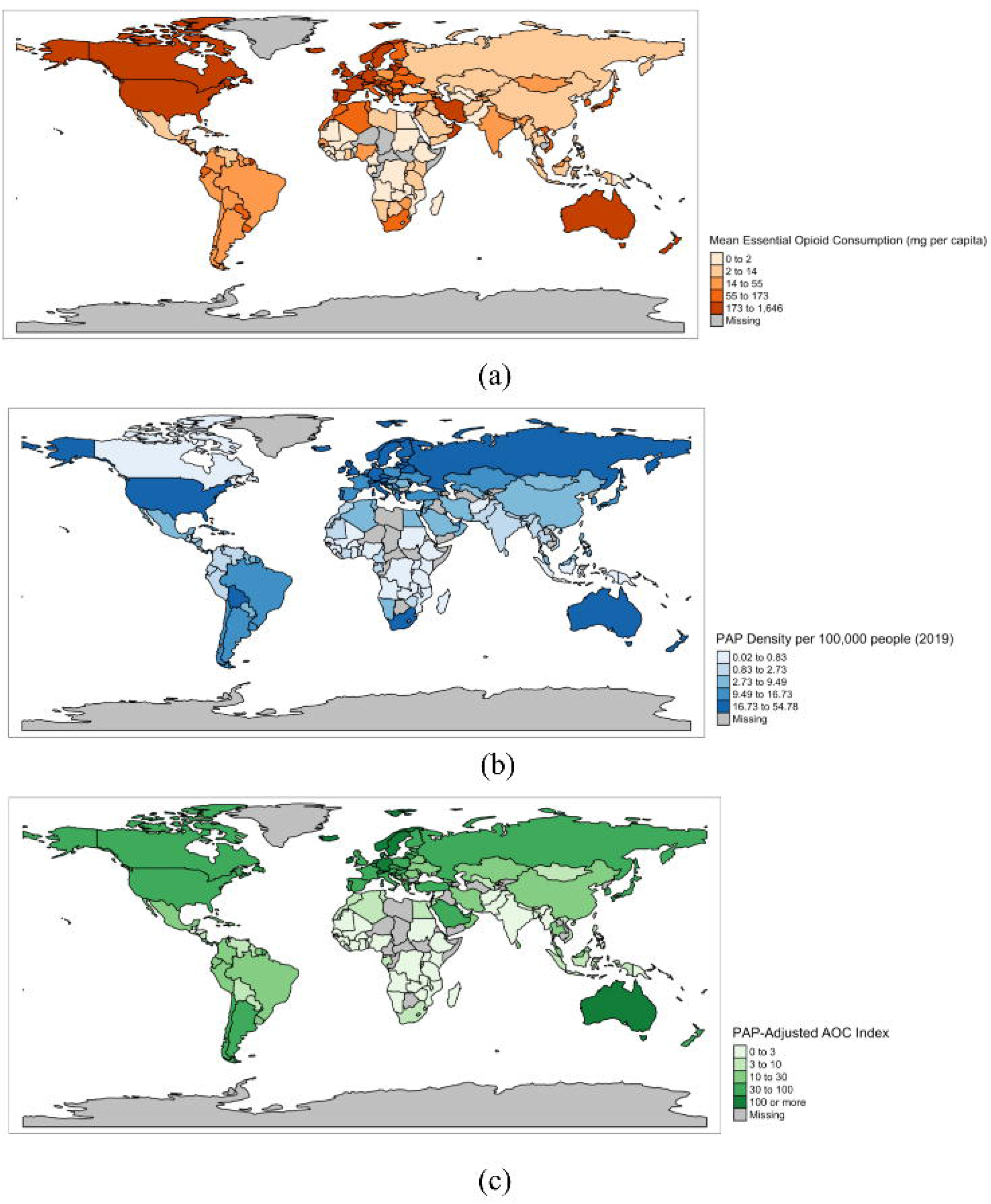

Globally, the median PAP density for 2019 was estimated to be 5.31 (IQR: 1.29-15.23) which ranged from 106.01 (95% UI: 154.15, 73.56) in Niue to 0.02 (95% UI: 0.01, 0.03) in Guinea **(Figure 1b)**. For validation of these projections, a strong positive correlation with (N=19, rho=0.900, p<0.001) was found with data for anesthetist densities for 2019 by Bouchard and colleagues.^16^

The PAP-adjusted AOC threshold was found to be 234.39 mg/capita. Using this threshold, the PAP-adjusted AOC index was calculated for 137 countries and territories as depicted in Figure 1c. Across countries, PAP-adjusted AOC values ranged from 129.14 for Switzerland to 0.23 for Mali **(Figure 1c)**. Of the 137 countries and territories merely 7.3% had a high, 23.36% had moderate, 21.17% had low, 16.79% had very low, and 31.39% of countries had extremely low PAP-adjusted AOC index values. About 5.97 billion people (77.5% of the global population) live in regions of low to extremely low PAP-adjusted AOC. Of those living in less than low adequacy regions, 3.4 billion (44.1%) are from LMICs and LICs and 2.98 billion (38.7%) belong to Southeast Asian and African WHO regions. A detailed account of the number of people under each adequacy category distributed across WBIGs and WHO regions is provided in **Table 1**.

**Table 1:**
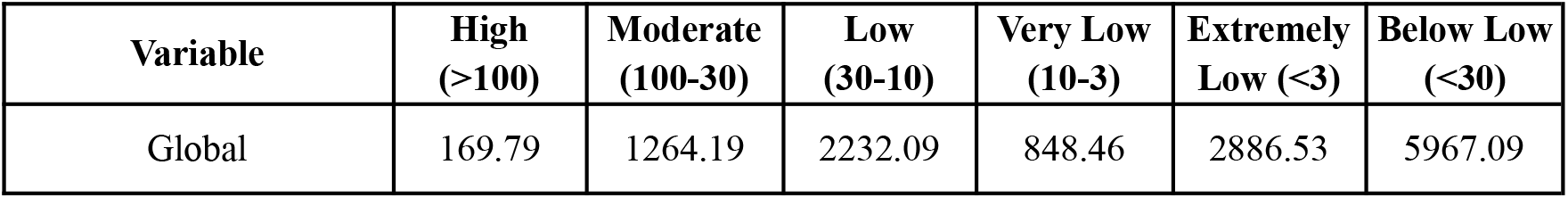
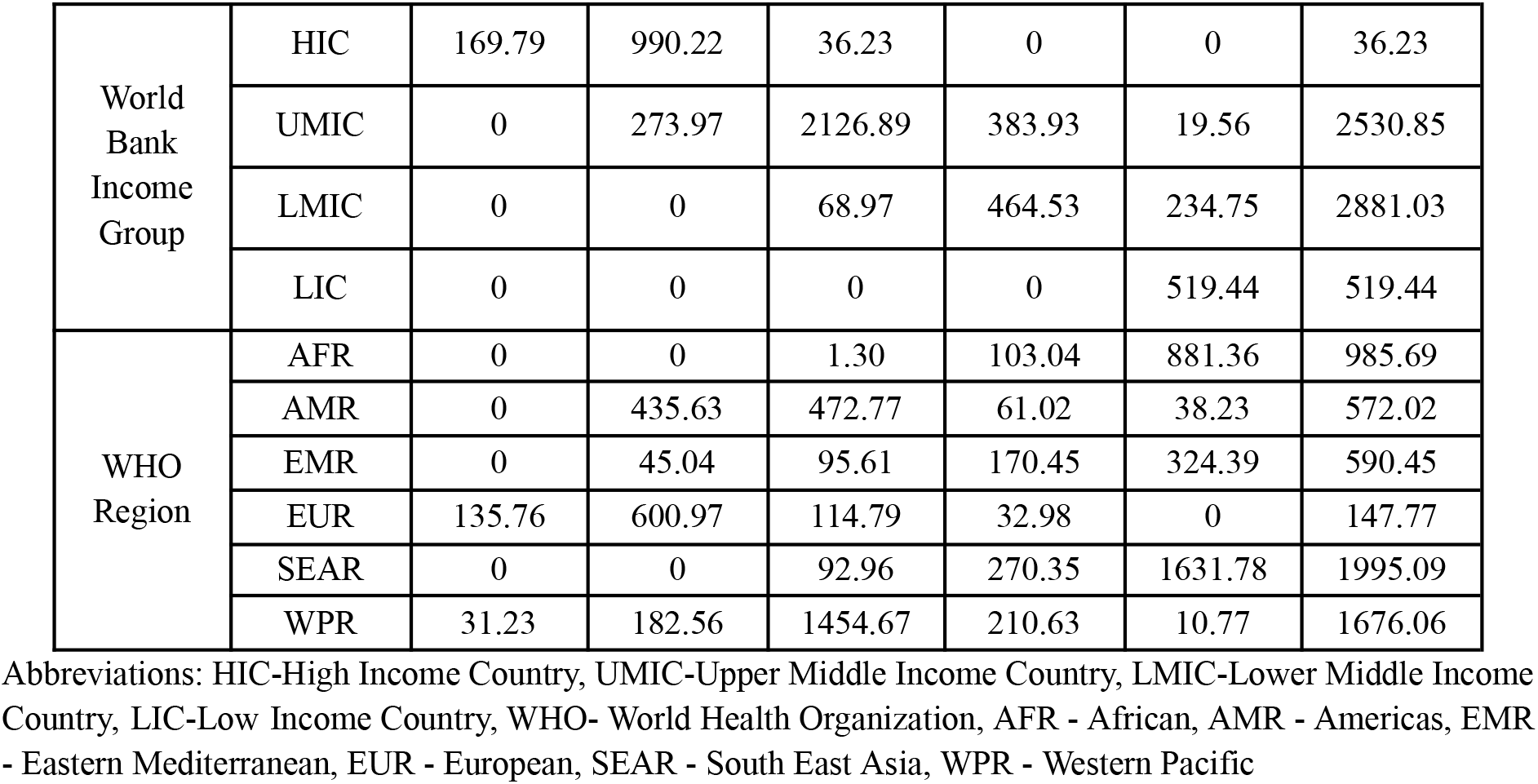
Number of people living across regions of different PAP-adjusted AOC categories (Population expression in millions)

| Variable | High (>100) | Moderate (100-30) | Low (30-10) | Very Low (10-3) | Extremely Low (<3) | Below Low (<30) |
| --- | --- | --- | --- | --- | --- | --- |
| Global | 169.79 | 1264.19 | 2232.09 | 848.46 | 2886.53 | 5967.09 |

|  |  |  |  |  |  |  |  |
| --- | --- | --- | --- | --- | --- | --- | --- |
| World Bank Income Group | HIC | 169.79 | 990.22 | 36.23 | 0 | 0 | 36.23 |
|  | UMIC | 0 | 273.97 | 2126.89 | 383.93 | 19.56 | 2530.85 |
|  | LMIC | 0 | 0 | 68.97 | 464.53 | 234.75 | 2881.03 |
|  | LIC | 0 | 0 | 0 | 0 | 519.44 | 519.44 |
| WHO Region | AFR | 0 | 0 | 1.30 | 103.04 | 881.36 | 985.69 |
|  | AMR | 0 | 435.63 | 472.77 | 61.02 | 38.23 | 572.02 |
|  | EMR | 0 | 45.04 | 95.61 | 170.45 | 324.39 | 590.45 |
|  | EUR | 135.76 | 600.97 | 114.79 | 32.98 | 0 | 147.77 |
|  | SEAR | 0 | 0 | 92.96 | 270.35 | 1631.78 | 1995.09 |
|  | WPR | 31.23 | 182.56 | 1454.67 | 210.63 | 10.77 | 1676.06 |

Abbreviations: HIC-High Income Country, UMIC-Upper Middle Income Country, LMIC-Lower Middle Income Country, LIC-Low Income Country, WHO- World Health Organization, AFR - African, AMR - Americas, EMR - Eastern Mediterranean, EUR - European, SEAR - South East Asia, WPR - Western Pacific

PAP-adjusted AOC values differed significantly across WHO regions (p<0.001). PAP-adjusted AOC values were higher for the European region compared to the Africa and SEARO regions **(Figure 2a)**. PAP-adjusted AOC also differed across country income groups (p=0.001). Post-hoc tests revealed that the values were significantly higher for high-income countries (HICs) compared to other groups **(Figure 2b)**.

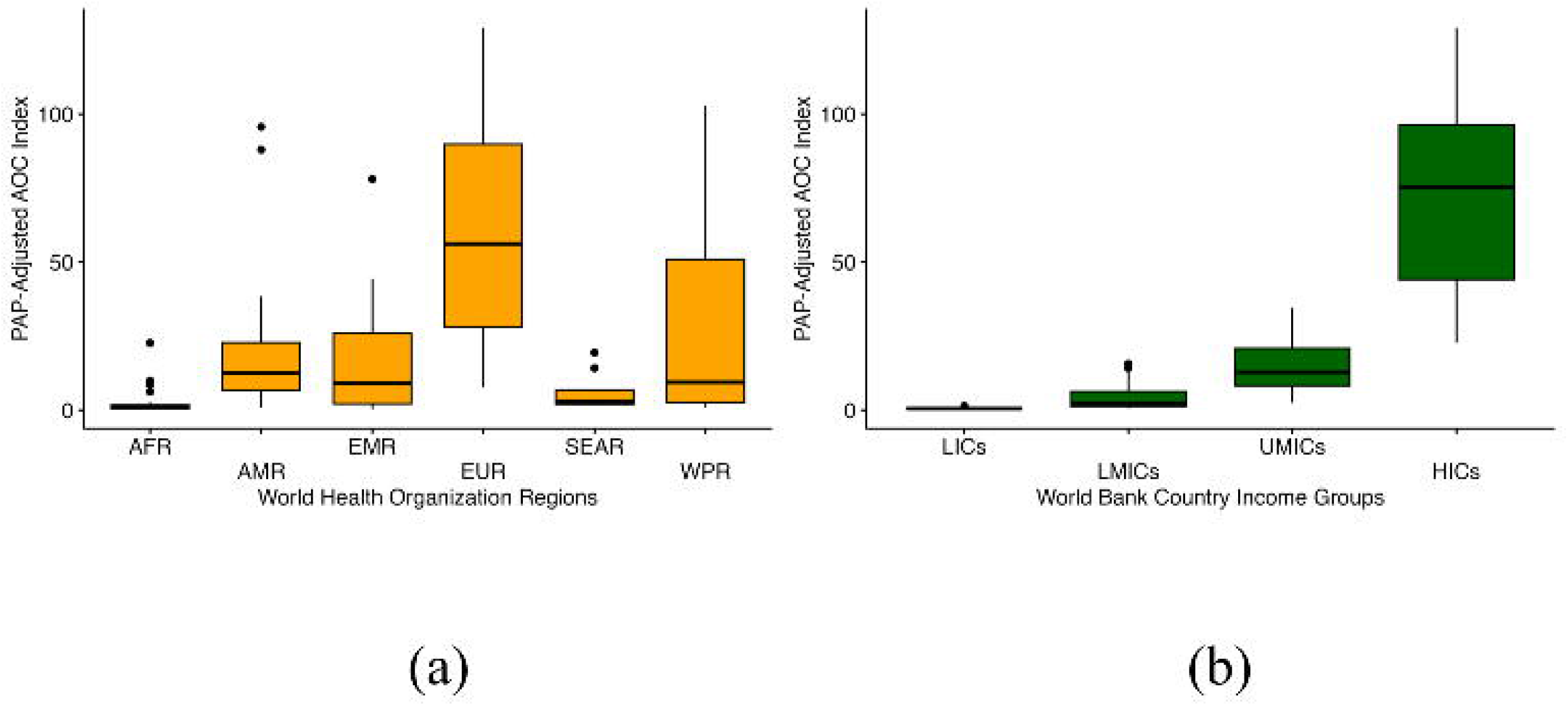

Based on calculations of the original AOC Index, the AOC threshold was 394.06 (mg/capita). On comparison of PAP-adjusted and original AOC indices, a non-linear moderate correlation (N=137, rho=0.821, p<0.001) was found using Spearman’s correlation **(Figure 3)**. The plot suggested that the values of these two variables were similar for middle values but were different at extremities. On further comparison, 56.93% (78/137) of the countries had a change in adequacy categories between original and PAP-adjusted AOC indices. **Figures 4a&b** depict the WBIGs and WHO region-wise change in AOC index categories for both indices.

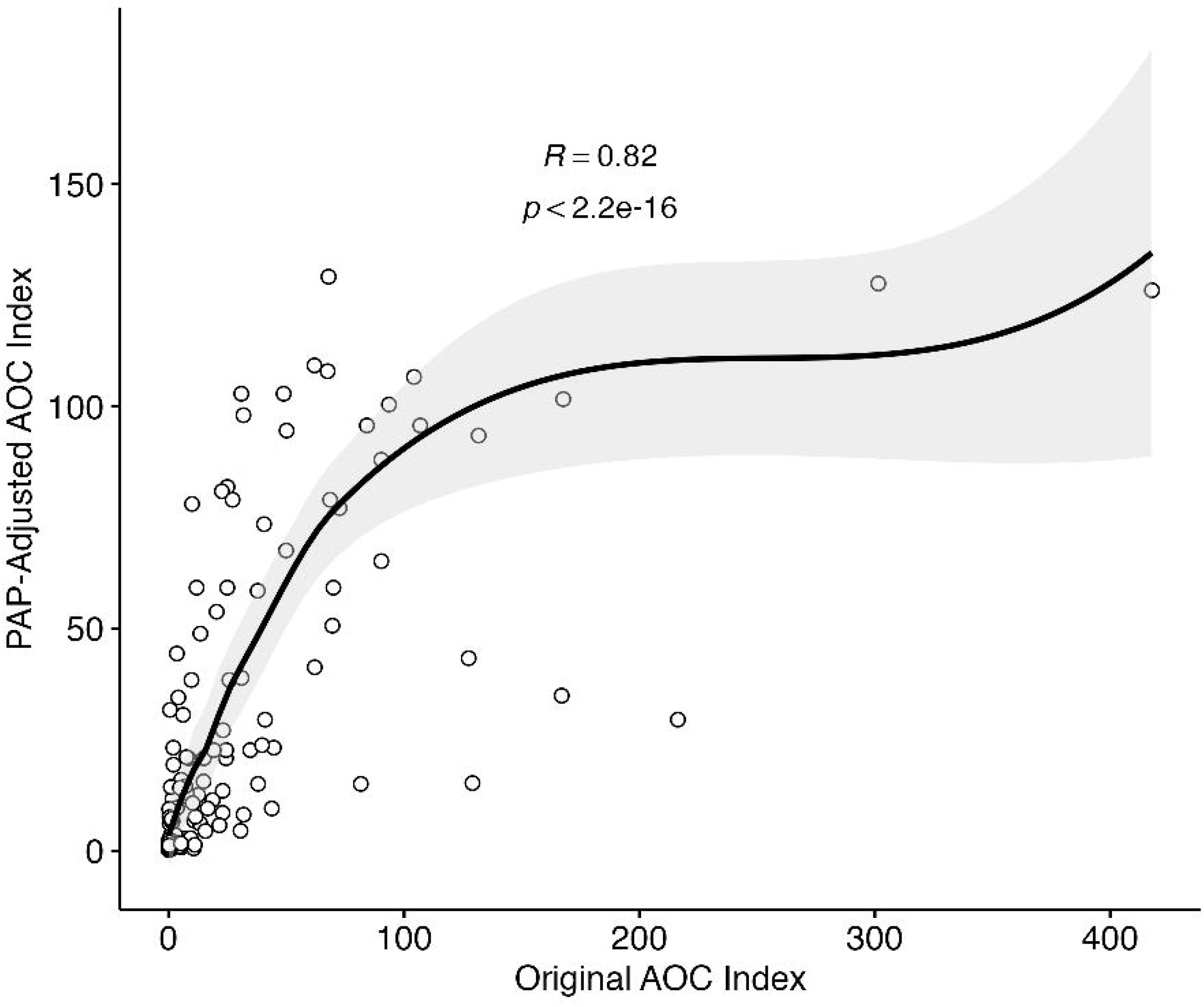

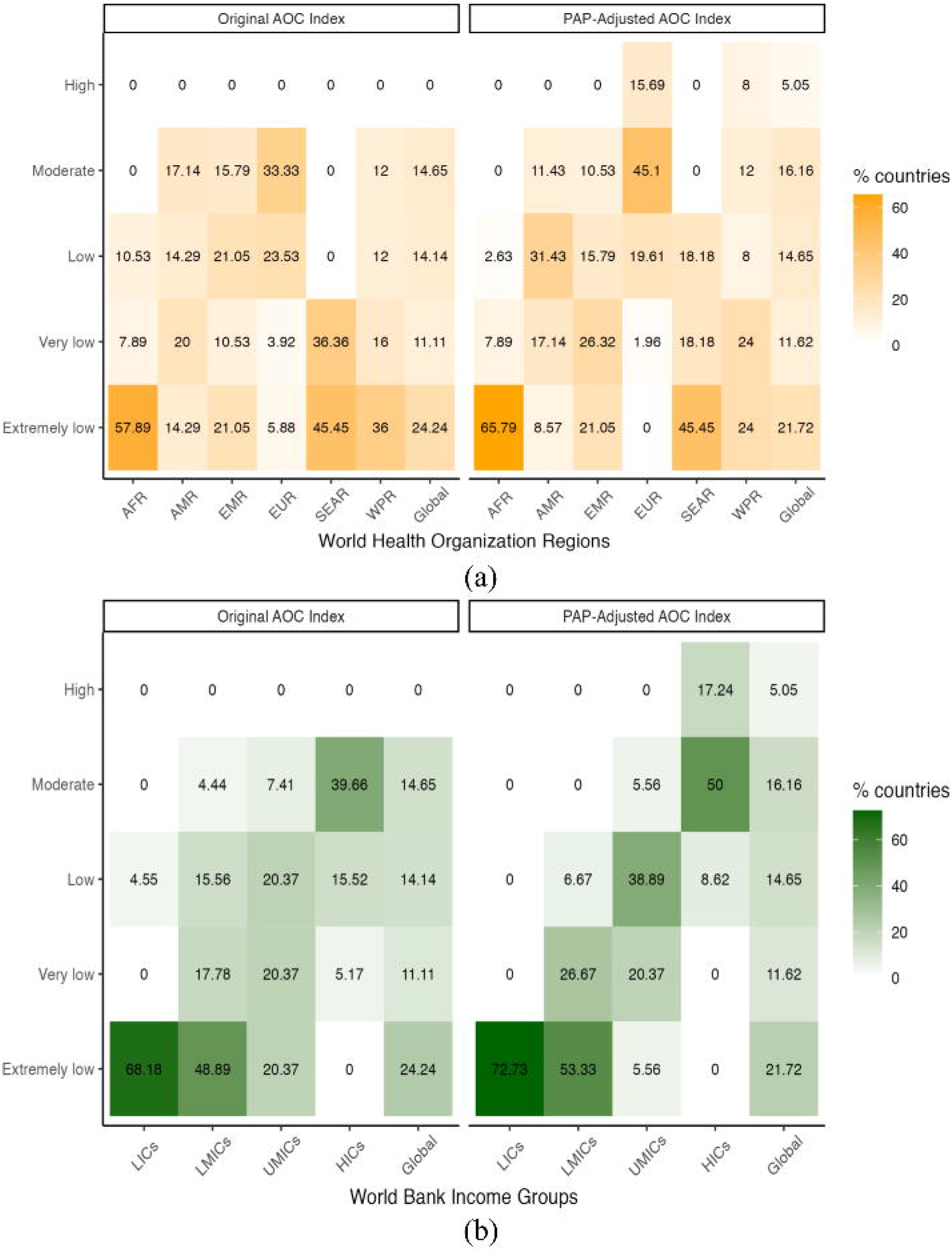

## Discussion

In this systematic global analysis of 137 countries, we estimate that 5.97 billion people i.e. 77.5% of the global population, live in regions of extremely low to low adequacy of essential opioid consumption. We find that most low- and middle-income countries situated in the global south lack adequate access to essential opioid analgesics. In 2010 using the ACM method, Duthey and Scholten found that 5.56 billion (79.3% of the global population) lived in countries with extremely low to low adequacy, which marginally increased to 5.96 billion (81.6% of the global population) in 2015. In terms of absolute values, an additional 400 million people in 2015 had inadequate access to opioid analgesics compared to 2010.^9^ According to our analysis, this number has decreased by 4.1 % compared to the last study.

During 2017-2021, the disparity in PAP-adjusted AOC was 562 times between the highest (Switzerland) and lowest (Mali) countries. In 2015, using the original AOC Index, Scholten et al. found a difference of approximately 44000 times between the highest (Germany) and lowest (Nigeria).^9^ Adjusting for PAP Density reduced the disparity of AOC between the highest and lowest countries considerably. Its advantage is that it normalizes the distribution of AOC by bringing higher and lower extremes closer to the mean adjusted consumption. Thus, the PAP-adjusted AOC accounts for opioid overconsumption values in developed countries. We have replaced “Adequate” with “High” while categorizing cut-offs for PAP-adjusted AOC, as the absence of an upper limit may lead to misinterpretation of the consumption levels which are much higher than 100 as adequate. This was done to account for countries with high AOC Index which may have over-prescription and over-consumption of opioids.^18^

Our methods differ from all of the previous studies in multiple ways. In terms of data sources, our data, although latest, is only limited to that available with the INCB. Some of the recent studies have estimated opioid analgesic consumption using the IQVIA MIDAS database, which estimates the consumption based on commercial sales and also accounts for the consumption of drugs that are not tracked by INCB.^7^ However data for less than 100 countries is available in that database, which limits the scope of global contextualization. We express our consumption data in mg/capita, which has been widely used in previous studies.^19^ However some studies have also used morphine milliequivalents (MMEs) to quantify mean opioid consumption.^9^ Studies have usually quantified the consumption of all types of prescription opioids, whose data was available at their respective data sources. These opioids were not only restricted to anesthesia and pain management but also included antitussive, antidiarrheals, etc. We have only included the consumption values for opioids which were included in the WHO Model List of Essential Medicines 2023 along with their analogues are used mainly for perioperative anesthesia and pain management. This makes our analysis more relevant to perioperative anesthesia and pain management indicators. We used PAP density as the health system indicator, for adjusted the AOC index. We used modeled data for the same as this data is not collected and is available annually. However, annual projections for physician densities are available based on which we devised and validated a method to not only project the annual PAP density but also to calculate the adjusted AOC of each year when new opioid consumption data is produced. This advantage of using PAP density as a health system indicator overcomes the limitations encountered by Scholten et al. concerning the limited availability and unreliability of disease morbidity data.^9^

However, beyond the healthcare factors, the supply and consumption of opioids are also governed by country-level regulations, policies, and protocols.^20^ Due to the well-documented risks associated with opioid medication including misuse, dependence, and the fear of being diverted from the domestic distribution channels to illicit circuits, in many countries, stringent legislative and regulatory interference along with cultural and local traditions are additional impediments to accessing opioid analgesics. Rendering opioids unavailable through restrictive legislation without acknowledging their rational medical use which is beneficial to patients in pain or with terminal illness may ultimately lead to avoidable, excruciating suffering.^21^

In the context of health planning and policy, PAP-adjusted AOC can be used as an indicator, especially in the fields of Global Surgical, Obstetric, Trauma, and Anesthesia (SOTA) care and palliative care to assess regional disparities in access to anesthesia and pain management. Inadequate access to analgesic opioids has direct consequences on perioperative care which is critical for attaining equitable SOTA care.^22^ National and sub-national mapping of PAP-adjusted AOC can help identify unmet needs and aid in drafting tailored healthcare plans addressing specific regional needs and challenges. AOC has already been recognized and used as an indicator in the South Asian Region for SOTA care situational analysis and healthcare planning.^23^

While novel, comprehensive, and valuable to global SOTA care research and policy agenda, our findings have certain limitations. Firstly, while calculating the five-year mean opioid consumption using the INCB data we acknowledge the data missingness for certain years and certain countries. Secondly, we have only accounted for one health system indicator, i.e., PAP density. Even though adjusting for PAP density normalizes the consumption value towards the mean, we could not define an upper limit for the overconsumption of opioids. Further research should be focused on adjusting the AOC Index based on other health system indicators like surgical rate and burden of disease for which more recent and reliable data needs to be produced. Additionally, defining an upper limit for the AOC Index is essential for setting benchmarks for overconsumption. Lastly, more global indicators for anesthesia and pain management should be explored which would aid in tracking its progress in global SOTA care development.

## Conclusion

We provide insights into the latest available data on prescription opioid consumption and its adequacy at the global, regional, and national levels using a new PAP-adjusted adequacy indicator. There is inadequate access to essential prescription opioid analgesics in most of the LMICs and LICs most of whom are situated in the global south, thus depriving close to 6 billion people of access to adequate perioperative care and pain management.

## Data Availability

All data used and produced in the manuscript can be accessed from Harvard Dataverse

https://doi.org/10.7910/DVN/F6RFMM

## Acknowledgments

None.

## Notes

**Competing interests:** The corresponding author Siddhesh Zadey is the Cofounding Director of the Association for Socially Applicable Research (ASAR). He also serves as the Permanent Council Member, The G4 Alliance Chair, SOTA Care in South Asia Working Group The G4 Alliance, and the Drafting Committee Member for Maharashtra State Mental Health Policy. The other authors declare no conflicts of interest.

### Competing Interest Statement

The corresponding author Siddhesh Zadey is the Cofounding Director of the Association for Socially Applicable Research (ASAR). He also serves as the Permanent Council Member, The G4 Alliance Chair, SOTA Care in South Asia Working Group The G4 Alliance, and the Drafting Committee Member for Maharashtra State Mental Health Policy. The other authors declare no conflicts of interest.

### Funding Statement

This study did not receive any funding

### Summary of Updates

The updated manuscript provides the latest available data for essential opioid analgesic consumption (190 countries) and adequacy (137 countries) between 2017-2021. We only include opioids and their analogs enlisted in the WHO list of Essential Drugs and use a modified AOC index which is adjusted for Physician Anesthesia Provider (PAP) density. This makes our index specific for anesthesia and perioperative pain management. Adjusting for PAP density normalizes the distribution of AOC by bringing higher and lower extremes closer to the mean adjusted consumption and thus accounts for opioid overconsumption values in developed countries.

