## Supplement 1 for "Adequacy of essential opioid analgesic consumption for anesthesia across 137 countries and territories from 2017 to 2021"

### Results

#### Descriptive Statistics

Descriptive Statistics

| | Log Opiod | Log Anathetist |
|---|---|---|
| Valid | 191 | 140 |
| Missing | 7 | 58 |
| Mean | 3.018 | 1.301 |
| Std. Deviation | 2.452 | 1.737 |
| Minimum | -5.498 | -3.876 |
| Maximum | 7.711 | 4.664 |

##### Distribution Plots

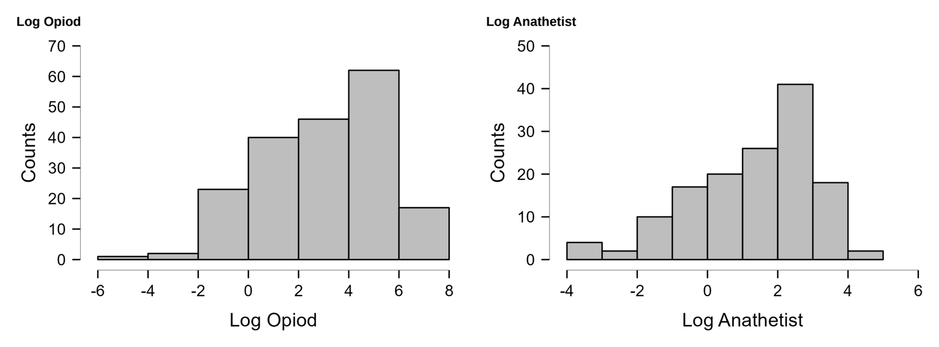


##### Correlation plot

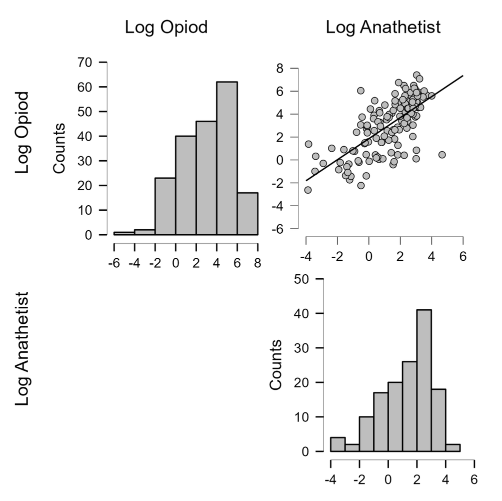


#### Linear Regression

Model Summary - Log Opiod

| Model | R | R² | Adjusted R² | RMSE | R² Change | F Change | df1 | df2 | p |
|---|---|---|---|---|---|---|---|---|---|
| $H_0$ | 0.000 | 0.000 | 0.000 | 2.319 | 0.000 | | 0 | 136 | |
| $H_1$ | 0.819 | 0.671 | 0.666 | 1.340 | 0.671 | 136.555 | 2 | 134 | < .001 |

ANOVA

| Model | | Sum of Squares | df | Mean Square | F | p |
|---|---|---|---|---|---|---|
| $H_1$ | Regression | 490.730 | 2 | 245.365 | 136.555 | < .001 |
| | Residual | 240.774 | 134 | 1.797 | | |
| | Total | 731.505 | 136 | | | |

*Note.* The intercept model is omitted, as no meaningful information can be shown.

Coefficients

| | | | | | | | 95% CI | | Collinearity Statistics | |
|---|---|---|---|---|---|---|---|---|---|---|
| Model | | Unstandardized | Standard Error | Standardized | t | p | Lower | Upper | Tolerance | VIF |
| $H_0$ | (Intercept) | 3.092 | 0.198 | | 15.607 | < .001 | 2.701 | 3.484 | | |
| $H_1$ | (Intercept) | -5.945 | 0.992 | | -5.992 | < .001 | -7.907 | -3.983 | | |
| | Log Anathetist | 0.092 | 0.128 | 0.068 | 0.716 | 0.475 | -0.162 | 0.345 | 0.269 | 3.716 |
| | HDI_2019 | 11.996 | 1.508 | 0.760 | 7.953 | < .001 | 9.012 | 14.979 | 0.269 | 3.716 |

Descriptives

| | N | Mean | SD | SE |
|---|---|---|---|---|
| Log Opiod | 137 | 3.092 | 2.319 | 0.198 |
| Log Anathetist | 137 | 1.278 | 1.731 | 0.148 |
| HDI_2019 | 137 | 0.744 | 0.147 | 0.013 |

Part And Partial Correlations

| Model | | Partial | Part |
|---|---|---|---|
| $H_1$ | Log Anathetist | 0.062 | 0.035 |
| | HDI_2019 | 0.566 | 0.394 |

*Note.* The intercept model is omitted, as no meaningful information can be shown.

Collinearity Diagnostics

| | | | | Variance Proportions | | |
|---|---|---|---|---|---|---|
| Model | Dimension | Eigenvalue | Condition Index | (Intercept) | Log Anathetist | HDI_2019 |
| $H_1$ | 1 | 2.541 | 1.000 | 0.002 | 0.018 | 0.002 |
| | 2 | 0.454 | 2.367 | 0.007 | 0.272 | 0.001 |
| | 3 | 0.006 | 21.258 | 0.991 | 0.710 | 0.997 |

*Note.* The intercept model is omitted, as no meaningful information can be shown.

Casewise Diagnostics

| Case Number | Std. Residual | Log Opiod | Predicted Value | Residual | Cook's Distance |
|---|---|---|---|---|---|
| 1 | 1.425 | 1.381 | -0.443 | 1.824 | 0.066 |
| 2 | 0.441 | 4.564 | 3.977 | 0.588 | 0.001 |
| 3 | 0.680 | 4.070 | 3.162 | 0.908 | 0.001 |
| 5 | -1.086 | -0.361 | 1.076 | -1.437 | 0.010 |
| 7 | -1.289 | 2.779 | 4.497 | -1.718 | 0.006 |
| 10 | 0.210 | 5.910 | 5.631 | 0.279 | 0.000 |
| 11 | 0.181 | 5.656 | 5.416 | 0.240 | 0.000 |
| 15 | -0.975 | 0.466 | 1.765 | -1.299 | 0.004 |
| 17 | 0.318 | 4.566 | 4.144 | 0.422 | 0.001 |
| 18 | -0.204 | 5.285 | 5.555 | -0.270 | 0.000 |
| 20 | -0.359 | -0.228 | 0.246 | -0.474 | 0.001 |
| 22 | -0.394 | 1.577 | 2.102 | -0.525 | 0.001 |
| 23 | 0.629 | 3.752 | 2.927 | 0.825 | 0.006 |
| 24 | 1.041 | 5.009 | 3.618 | 1.390 | 0.003 |
| 26 | -0.036 | 3.421 | 3.468 | -0.048 | 0.000 |
| 30 | -0.834 | -1.732 | -0.635 | -1.097 | 0.009 |
| 34 | 0.643 | 6.043 | 5.248 | 0.795 | 0.024 |
| 36 | -0.722 | 3.641 | 4.604 | -0.962 | 0.002 |
| 37 | -1.137 | 1.827 | 3.346 | -1.519 | 0.003 |
| 41 | 0.453 | 3.900 | 3.298 | 0.602 | 0.001 |
| 44 | 0.954 | 5.170 | 3.914 | 1.257 | 0.011 |
| 45 | -0.482 | 0.102 | 0.734 | -0.632 | 0.003 |
| 46 | -0.032 | 4.621 | 4.664 | -0.043 | 0.000 |
| 47 | -0.547 | 3.039 | 3.765 | -0.726 | 0.002 |
| 49 | 0.418 | 5.619 | 5.063 | 0.556 | 0.001 |
| 50 | -0.939 | 3.845 | 5.094 | -1.249 | 0.005 |
| 52 | -0.386 | -0.848 | -0.339 | -0.509 | 0.002 |
| 53 | 0.220 | 6.018 | 5.726 | 0.292 | 0.000 |
| 55 | -0.063 | 3.412 | 3.496 | -0.084 | 0.000 |
| 56 | 0.801 | 4.300 | 3.233 | 1.067 | 0.003 |
| 57 | -1.945 | 0.448 | 3.040 | -2.592 | 0.014 |
| 58 | 1.835 | 4.790 | 2.342 | 2.448 | 0.011 |
| 60 | -0.062 | 5.005 | 5.087 | -0.083 | 0.000 |
| 61 | 0.015 | 1.427 | 1.407 | 0.020 | 0.000 |
| 62 | -0.051 | -0.303 | -0.237 | -0.066 | 0.000 |
| 64 | -0.425 | 2.543 | 3.111 | -0.568 | 0.000 |
| 65 | -0.577 | 4.830 | 5.594 | -0.765 | 0.002 |
| 66 | 0.542 | 5.875 | 5.154 | 0.721 | 0.002 |
| 68 | -1.619 | 0.439 | 2.598 | -2.159 | 0.009 |
| 69 | 0.050 | 4.077 | 4.010 | 0.067 | 0.000 |
| 70 | -0.185 | 5.497 | 5.743 | -0.246 | 0.000 |
| 71 | -0.594 | 0.753 | 1.541 | -0.788 | 0.003 |
| 73 | -0.426 | 4.386 | 4.952 | -0.566 | 0.001 |
| 75 | 1.096 | 3.281 | 1.820 | 1.460 | 0.005 |
| 76 | -1.502 | -2.625 | -0.698 | -1.927 | 0.069 |
| 77 | 0.975 | 3.968 | 2.667 | 1.301 | 0.003 |
| 78 | -0.520 | -0.117 | 0.571 | -0.688 | 0.002 |
| 79 | -0.135 | 1.532 | 1.712 | -0.180 | 0.000 |
| 80 | 1.498 | 6.489 | 4.492 | 1.997 | 0.009 |
| 81 | 1.175 | 7.406 | 5.850 | 1.556 | 0.012 |
| 82 | 1.337 | 3.602 | 1.820 | 1.782 | 0.006 |
| 83 | -0.848 | 1.497 | 2.622 | -1.125 | 0.005 |
| 84 | 1.628 | 5.774 | 3.601 | 2.173 | 0.007 |
| 86 | 0.661 | 6.493 | 5.616 | 0.877 | 0.003 |
| 87 | 0.216 | 5.600 | 5.314 | 0.286 | 0.000 |
| 88 | -0.399 | 4.586 | 5.117 | -0.531 | 0.001 |
| 90 | -0.573 | 4.588 | 5.346 | -0.759 | 0.003 |
| 91 | 0.641 | 3.810 | 2.957 | 0.853 | 0.002 |
| 92 | -1.573 | 2.042 | 4.136 | -2.094 | 0.012 |
| 93 | 1.103 | 2.418 | 0.953 | 1.465 | 0.007 |
| 94 | -0.841 | 0.630 | 1.743 | -1.113 | 0.007 |
| 95 | 0.544 | 5.084 | 4.358 | 0.725 | 0.001 |
| 98 | 1.087 | 6.219 | 4.772 | 1.447 | 0.006 |
| 99 | 0.733 | 4.180 | 3.205 | 0.975 | 0.003 |
| 103 | 0.512 | 5.613 | 4.933 | 0.681 | 0.001 |
| 105 | -0.485 | -0.596 | 0.045 | -0.641 | 0.002 |
| 106 | 0.271 | 0.320 | -0.029 | 0.349 | 0.002 |
| 107 | -0.272 | 3.512 | 3.875 | -0.363 | 0.000 |
| 109 | -0.441 | -1.433 | -0.856 | -0.578 | 0.003 |
| 110 | -0.178 | 5.073 | 5.309 | -0.236 | 0.000 |
| 112 | -1.645 | -1.390 | 0.794 | -2.183 | 0.018 |
| 113 | 0.244 | 4.323 | 3.998 | 0.325 | 0.000 |
| 114 | -1.715 | 1.279 | 3.569 | -2.290 | 0.008 |
| 115 | -1.666 | -0.411 | 1.790 | -2.201 | 0.027 |
| 116 | -0.409 | 2.636 | 3.182 | -0.546 | 0.001 |
| 119 | 1.362 | 4.117 | 2.299 | 1.817 | 0.005 |
| 120 | -0.570 | -1.361 | -0.610 | -0.751 | 0.004 |
| 121 | 0.973 | 2.553 | 1.260 | 1.293 | 0.005 |
| 122 | 0.580 | 2.630 | 1.863 | 0.767 | 0.003 |
| 123 | 1.017 | 2.726 | 1.371 | 1.354 | 0.005 |
| 124 | -0.610 | 4.801 | 5.608 | -0.807 | 0.003 |
| 126 | 0.172 | 5.805 | 5.576 | 0.229 | 0.000 |
| 127 | 0.188 | 2.404 | 2.155 | 0.249 | 0.000 |
| 128 | 1.962 | 3.061 | 0.461 | 2.600 | 0.029 |
| 131 | 1.919 | 6.232 | 3.674 | 2.557 | 0.015 |
| 132 | 0.907 | 7.080 | 5.879 | 1.201 | 0.007 |
| 133 | 1.842 | 6.748 | 4.293 | 2.455 | 0.013 |
| 134 | -0.303 | 0.257 | 0.656 | -0.399 | 0.001 |
| 136 | 0.637 | 4.917 | 4.067 | 0.849 | 0.001 |
| 137 | -2.229 | -2.233 | 0.727 | -2.960 | 0.032 |
| 138 | 1.381 | 4.831 | 2.990 | 1.842 | 0.006 |
| 139 | -0.101 | 3.331 | 3.465 | -0.134 | 0.000 |
| 140 | -0.791 | 1.728 | 2.784 | -1.055 | 0.002 |
| 141 | -0.682 | 3.973 | 4.881 | -0.908 | 0.002 |
| 142 | 0.582 | 5.499 | 4.725 | 0.775 | 0.002 |
| 144 | -0.651 | 4.490 | 5.353 | -0.863 | 0.003 |
| 145 | 0.703 | 4.505 | 3.570 | 0.935 | 0.003 |
| 146 | 0.218 | 4.510 | 4.219 | 0.291 | 0.000 |
| 147 | -2.726 | 0.844 | 4.469 | -3.625 | 0.039 |
| 148 | 0.323 | 0.795 | 0.367 | 0.428 | 0.001 |
| 153 | -0.581 | 1.900 | 2.675 | -0.775 | 0.001 |
| 154 | -1.551 | 2.622 | 4.679 | -2.057 | 0.017 |
| 155 | 2.701 | 3.737 | 0.169 | 3.567 | 0.072 |
| 156 | -0.458 | 3.399 | 4.010 | -0.611 | 0.001 |
| 158 | -0.380 | -0.993 | -0.501 | -0.492 | 0.004 |
| 159 | -0.231 | 5.260 | 5.565 | -0.305 | 0.001 |
| 161 | 0.116 | 4.804 | 4.650 | 0.154 | 0.000 |
| 162 | -0.527 | 4.673 | 5.373 | -0.700 | 0.002 |
| 163 | 1.566 | 2.927 | 0.848 | 2.079 | 0.016 |
| 164 | 1.031 | 4.503 | 3.143 | 1.360 | 0.011 |
| 165 | 0.057 | 5.281 | 5.205 | 0.076 | 0.000 |
| 166 | -0.387 | 2.940 | 3.455 | -0.515 | 0.001 |
| 167 | -0.274 | -0.199 | 0.163 | -0.363 | 0.001 |
| 168 | 0.348 | 3.694 | 3.229 | 0.465 | 0.000 |
| 169 | -0.087 | 5.584 | 5.699 | -0.115 | 0.000 |
| 170 | -0.281 | 5.590 | 5.962 | -0.372 | 0.001 |
| 171 | 1.982 | 3.776 | 1.167 | 2.608 | 0.049 |
| 173 | -1.288 | 2.077 | 3.789 | -1.712 | 0.009 |
| 174 | 0.717 | 2.366 | 1.412 | 0.954 | 0.002 |
| 175 | -1.091 | -1.094 | 0.350 | -1.444 | 0.010 |
| 176 | -2.456 | -0.151 | 3.127 | -3.278 | 0.017 |
| 178 | 1.503 | 5.152 | 3.145 | 2.007 | 0.007 |
| 179 | -0.885 | 3.181 | 4.362 | -1.180 | 0.003 |
| 182 | -1.436 | 0.106 | 1.948 | -1.843 | 0.063 |
| 183 | 0.525 | 0.901 | 0.207 | 0.694 | 0.003 |
| 184 | 0.252 | 4.066 | 3.731 | 0.336 | 0.000 |
| 185 | -1.241 | 3.668 | 5.314 | -1.646 | 0.011 |
| 186 | 0.539 | 6.252 | 5.536 | 0.716 | 0.002 |
| 187 | 0.459 | 1.014 | 0.412 | 0.601 | 0.003 |
| 188 | 0.291 | 5.875 | 5.489 | 0.386 | 0.001 |
| 189 | 0.672 | 5.052 | 4.158 | 0.894 | 0.002 |
| 190 | -1.929 | 0.410 | 2.970 | -2.560 | 0.025 |
| 191 | -0.939 | 0.179 | 1.428 | -1.249 | 0.005 |
| 192 | -0.935 | 1.529 | 2.777 | -1.249 | 0.002 |
| 193 | 1.461 | 4.442 | 2.497 | 1.945 | 0.010 |
| 196 | -0.728 | -0.077 | 0.890 | -0.967 | 0.003 |
| 197 | 1.333 | 3.028 | 1.254 | 1.774 | 0.009 |

Residuals Statistics

| | Minimum | Maximum | Mean | SD | N |
|---|---|---|---|---|---|
| Predicted Value | -0.856 | 5.962 | 3.092 | 1.900 | 137 |
| Residual | -3.625 | 3.567 | $4.296\times10^{-17}$ | 1.331 | 137 |
| Std. Predicted Value | -2.078 | 1.511 | $4.748\times10^{-19}$ | 1.000 | 137 |
| Std. Residual | -2.726 | 2.701 | $-1.792\times10^{-4}$ | 1.003 | 137 |

##### Residuals vs. Dependent

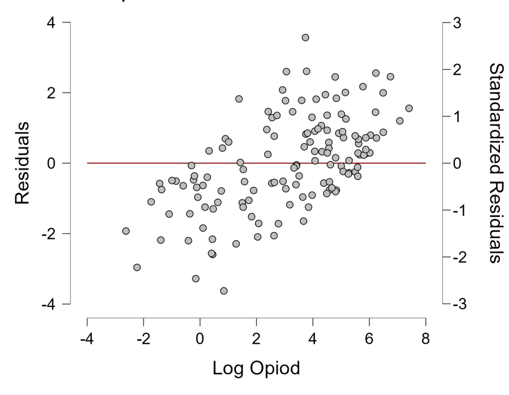


##### Residuals vs. Covariates

Residuals vs. Log Anathetist

Residuals vs. HDI_2019

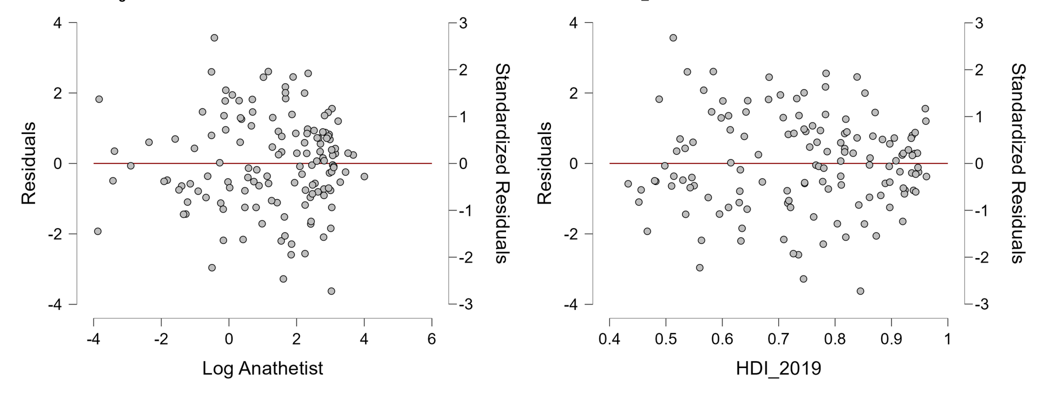


Residuals vs. Predicted

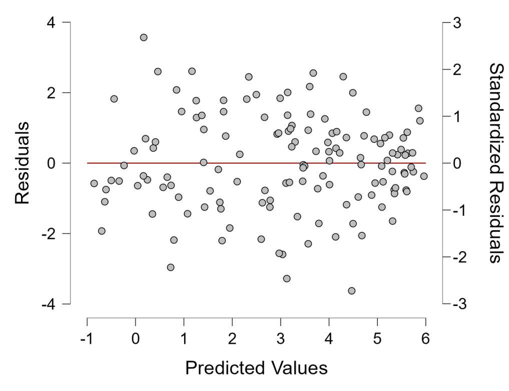


Standardized Residuals Histogram

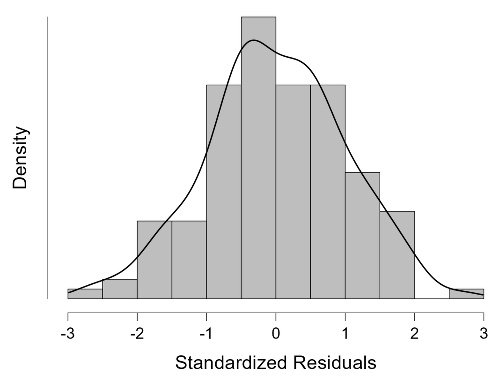


Q-Q Plot Standardized Residuals

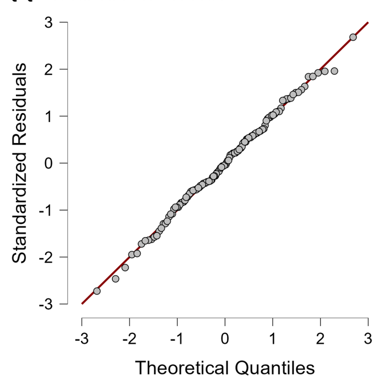


##### Partial Regression Plots

Log Opiod vs. Log Anathetist

Log Opiod vs. HDI_2019

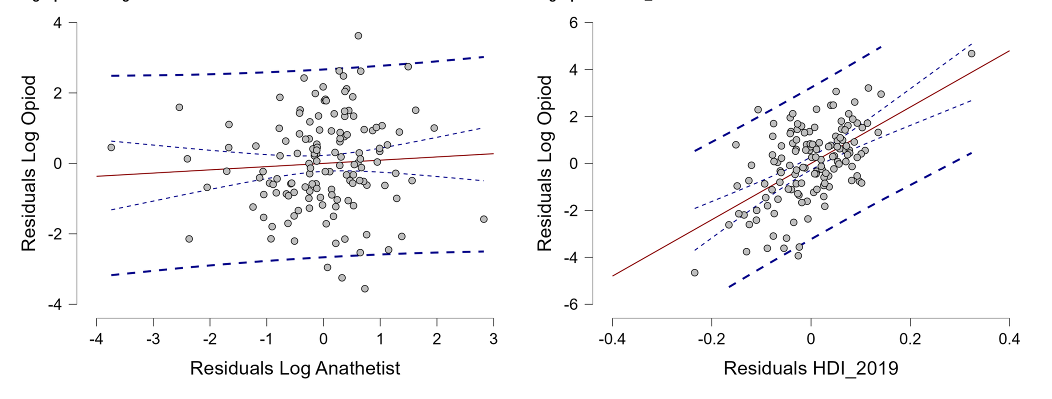


##### Marginal Effects Plots

Marginal effect of Log Anathetist on Log Opiod

Marginal effect of HDI_2019 on Log Opiod

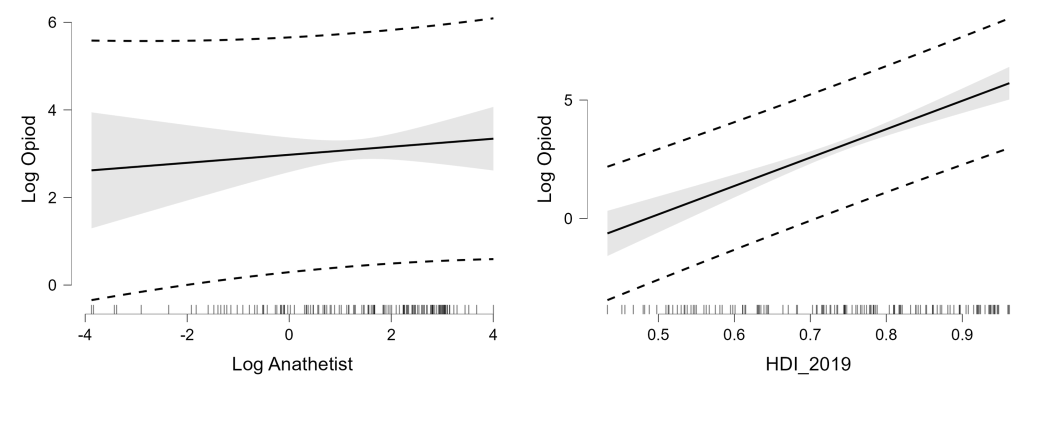


#### Generalized Linear Model

Model Summary - Log Opiod

| Model | Deviance | AIC | BIC | df | Χ² | p |
|---|---|---|---|---|---|---|
| $H_0$ | 731.505 | 622.281 | 628.121 | 136 | | |
| $H_1$ | 240.774 | 474.041 | 485.721 | 134 | 490.730 | < .001 |

Coefficients

| | Estimate | Standard Error | t | p |
|---|---|---|---|---|
| (Intercept) | -5.945 | 0.992 | -5.992 | < .001 |
| Log Anathetist | 0.092 | 0.128 | 0.716 | 0.475 |
| HDI_2019 | 11.996 | 1.508 | 7.953 | < .001 |

##### Estimated Marginal Means and Contrast Analysis

Estimated Marginal Means

| HDI_2019 | Estimate | SE |
|---|---|---|
| 0.597 | 1.330 | 0.249 |
| 0.744 | 3.092 | 0.115 |
| 0.891 | 4.855 | 0.249 |

*Note.* Results are on the response scale.

#### Descriptive Statistics

Descriptive Statistics

| | . |
|---|---|
| Valid | . |
| Missing | . |
| Mean | . |
| Std. Deviation | . |
| Minimum | . |
| Maximum | . |

#### Correlation

Spearman's Correlations

| Variable | | Bochard A Density per 100,000 | Anathetist Density 2019 |
|---|---|---|---|
| 1. Bochard A Density per 100,000 | Spearman's rho | — | |
| | p-value | — | |
| 2. Anathetist Density 2019 | Spearman's rho | 0.900 | — |
| | p-value | < .001 | — |

##### Assumption checks

Shapiro-Wilk Test for Multivariate Normality

| Shapiro-Wilk | p |
|---|---|
| 0.663 | < .001 |

Shapiro-Wilk Test for Bivariate Normality

| | | | Shapiro-Wilk | p |
|---|---|---|---|---|
| Bochard A Density per 100,000 | - | Anathetist Density 2019 | 0.663 | < .001 |

##### Correlation plot

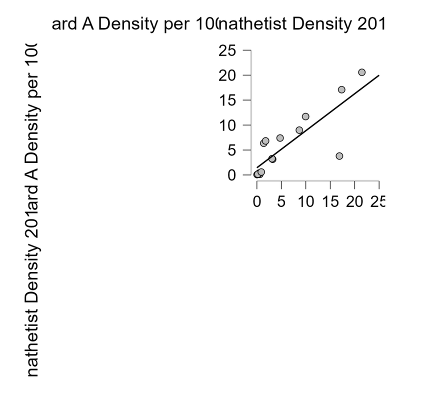
